# Interpretable machine-learning model for cataract associated factors identifying in patients with high myopia

**DOI:** 10.64898/2026.02.25.26347145

**Authors:** Kaimeng Su, Qihao Duan, Wenwen He, Benjamin Wild, Roland Eils, Lei Gu, Xiangjia Zhu

## Abstract

**Purpose:** To systematically evaluate ocular biometric and systemic laboratory factors associated with cataract in highly myopic eyes and to characterize potential nonlinear associations using an interpretable machine learning approach, thereby providing deeper mechanistic insights into the pathogenesis of highly myopic cataract.

**Design:** A cross-sectional study encompassed 770 eyes of 594 patients with high myopia from Eye & ENT Hospital of Fudan University.

**Subjects:** The non-cataract control group included 458 eyes while the cataract group contained 312 eyes.

**Methods:** Demographic traits, ocular biometric and systemic laboratory factors were gathered while features with over 30% of missing data were excluded. Composite indices were obtained through calculation. Multiple machine learning models were compared to investigate the association between features and highly myopic cataract, and the random forest (RF) model was chosen and fine-tuned. Feature selection was carried out by means of Shapley additive explanations (SHAP) and non-linear relationships were probed using SHAP dependence diagrams and confirmed with partial dependence plots.

**Main Outcome Measures:** (1) The Area Under the Curve (AUC) and other metrics of multiple machine learning models; (2) Top feature importance of the final simplified RF model; (3) Overall trends between features and highly myopic cataract; (4) Potential inflection points of top continuous features.

**Results:** A simplified fine-tuned RF model with 17 features reached stable discriminative performance, with a mean AUC of 0.762 (95%CI: [0.731, 0.794]) among 10 independent testing sets. Age and axial length (AL) turned out to be the most influential features which had non-linear relationships highly myopic cataract, with an inflection point seen around 65.75 (95%CI: [63.72, 67.79]) years for age and 30.55 (95% CI: [29.22, 31.88]) mm for axial length respectively, while the ratio of anterior chamber depth to axial length (ACD/AL) was associated with highly-myopic cataract in a U-shape. Ocular biometric factors were more strongly related to highly myopic cataract than systemic laboratory factors.

**Conclusions:** Ocular biometric factors, especially age, AL, and composite indices like ACD/AL, have strong and non-linear connections with highly myopic cataract. These results emphasize the significance of ocular structural arrangement in cataract within highly myopic eyes and indicate that interpretable data-driven methods could offer clinically relevant understandings regarding its phenotypic description.

## Introduction

Projections indicate that by 2050 almost half of the world’s population might be myopia and around 10% could have high myopia, which leads to an expanding group of people being at risk of vision-threatening complications[1]. In line with this tendency, the International Myopia Institute (IMI) pointed out that the public health burden of myopia is increasingly caused by its complications, stressing the necessity to alter clinical priorities towards preventing long-term illness[2].

Of these complications, cataract is the most common, particularly in individuals with longer axial length or higher degrees of myopia, which has long been a major cause of global visual impairment and blindness[3–8]. Prior work has shown that myopia and AL are associated with cataract[9]. Although high myopia is characterized by abnormal AL elongation and remodeling of ocular geometry[10], it remains unclear how ocular biometric parameters indicate cataract in highly myopic eyes. Notably, beyond AL, overall eye shape can be quantified using multidimensional biometric measures and composite indices, which may better capture the anatomic context relevant to normal lens function. However, the specific contributions of these factors to highly myopic cataract remain insufficiently defined. In addition, prior studies have suggested threshold-like effects in the relationship between myopia, AL and cataract, implying potential nonlinearity[11]. Whether similar nonlinear associations exist between AL and cataract specifically within highly myopic eyes, and whether inflection points can be identified, remains to be determined.

Additionally, high myopia has been discussed in the context of immune and inflammatory pathways, with some studies showing that there are connections between myopia and circulating immune cell profiles or systemic inflammatory markers[12, 13]. However, it’s still unclear whether common lab tests like hematologic, biochemical, coagulation, and immunologic evaluations offer any additional insights into cataracts in individuals with high myopia.

Another major deficiency is that previous research frequently concentrated on whether variables were related to cataract instead of how these relationships acted across the predictor range. Nonlinear patterns as well as inflection points might be clinically useful, perhaps indicating stage-dependent changes in ocular biomechanics, zonular tension, or metabolic balance, but traditional modeling methods could overlook such patterns when linearity was presumed or interactions had to be pre-determined.

In this research, we methodically investigated age, sex, ocular biometric factors and systemic laboratory factors related to cataracts in eyes with high myopia. By employing an interpretable machine-learning method with strict cross-validation and multiple rounds of training-testing splits, we endeavored to spot a concise collection of major associated features and describe the nature of these associations, like non-linear patterns and possible turning points, thus offering clinically comprehensible understandings regarding cataracts in highly myopic eyes.

## Methods

### Ethics approval and consent to participate

The research plan was approved by the Ethics Committee of the Eye & ENT Hospital affiliated with Fudan University (Shanghai, China). All steps regarding human participants were carried out in line with the ethical guidelines of the Declaration of Helsinki as well as the approved research plan. Written agreement with informed consent was obtained from every participant before they took part in the study. Moreover, this study had been registered on ClinicalTrials.gov (ID: NCT03062085).

### Study Design and Participants

The overall study design and analysis workflow are summarized in **Figure 1**.

**Figure 1.**
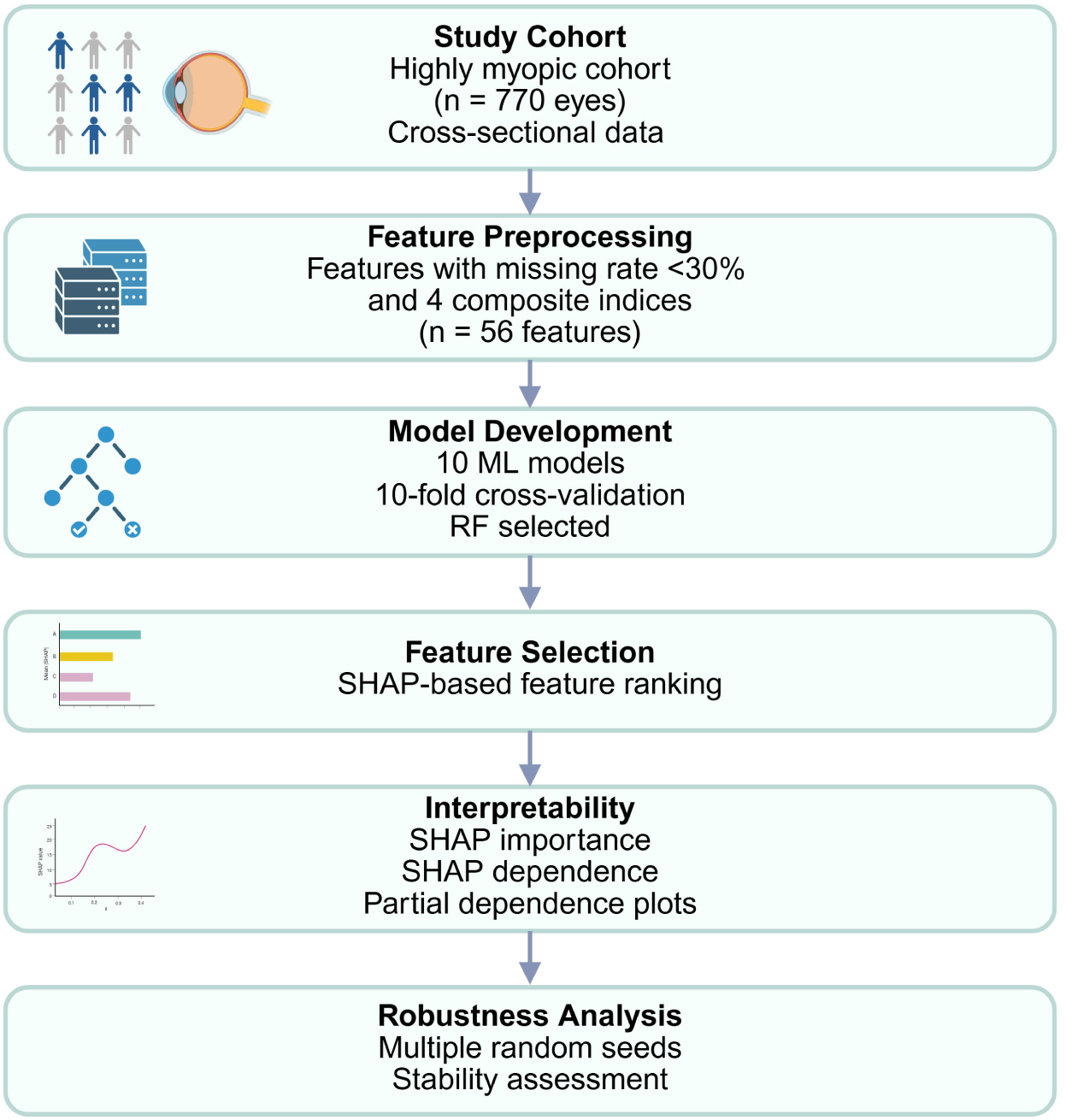
Study design and analysis workflow. In this study, a cross-sectional group of eyes with high myopia was incorporated, characteristics with a missing proportion of under 30% were kept for examination, the data set was randomly divided into training and testing parts in a 9:1 ratio and numerous machine learning (ML) models were evaluated through 10-fold cross-validation. A random forest (RF) model was chosen considering its overall discriminative ability. The significance of features and the model’s interpretability were measured by means of Shapley additive explanations (SHAP) and the non-linear impacts of crucial features were further explored via SHAP dependence plots and partial dependence plots, the model’s robustness was assessed across several random data divisions. Abbreviations: ML, machine learning; RF, random forest; SHAP, Shapley additive explanations.

This cross-sectional study utilized clinical data from the Fudan University Eye & ENT Hospital spanning from 2016 to 2025 and a total of 770 highly myopic eyes were involved, with 458 eyes free of cataract and 312 eyes having been diagnosed with cataract by two professional ophthalmologists, and each eye was regarded as an individual analytical unit.

Demographic traits such as age and sex were collected at the time of clinical presentation. Ocular biometric factors encompassed AL, keratometry figures (K1 and K2), anterior chamber depth (ACD), central corneal thickness (CCT), and corneal endothelial cell count (CECs), which were obtained using optical biometers (IOL-Master 500 and IOL-Master 700, Carl Zeiss, Germany).

Systemic laboratory factors encompassed hematological, coagulation, biochemical, and immunological indices, including blood cell counts and differentials, liver and renal function markers, coagulation parameters, and infection-related serology.

The data distribution of the disease outcome and all included features was summarized for the training set and the testing set respectively. Welch’s t-test was applied to continuous variables among the training and testing sets, while the chi-square test was applied to categorical variables.

### Feature Collection and Preprocessing

Initially, a full range of clinical and ocular biometric variables was gathered and features with a missing-data rate of more than 30% were removed to guarantee data quality and model stability. After this filtering process, a total of 52 features remained for the ensuing analyses.

Besides the initial measurements, four combined indices were developed to better capture ocular anatomic coupling and proportional relationships that may not be fully represented by single parameters, including average keratometry (calculated as (K1 + K2) / 2), the proportion of anterior chamber depth to axial length (denoted as ACD / AL), absolute keratometric astigmatism (which is the absolute value of K1 - K2), and the multiplication result of keratometry figures (K1 multiplied by K2). A comprehensive list along with definitions of all the features involved was offered in ***Supplementary Table S1***.

The dataset got randomly split into training and testing sets in a 9:1 ratio using a fixed random seed, with distributions of cataract diagnosis in training and testing sets close to the original. For the purpose of preventing information seepage, missing values were filled in separately inside the training and testing sets through the *miceforest* package (version 5.6.0, Python 3.9) which carried out multiple imputation by chained equations depending on random forests and when cross-validation was taking place[14, 15]. Specifically, the imputation models were fit on the training data only and then applied to impute the corresponding missing values in the testing data. Imputation was done independently within every fold.

### Model Development and Comparison

A number of machine learning (ML) algorithms were examined, such as logistic regression, Lasso regression, elastic net, Gaussian naive Bayes, support vector machine, random forest (RF), gradient boosting, light gradient boosting machine (LightGBM), extreme gradient boosting (XGBoost), and TabPFN. Model’s performance was evaluated by means of 10-fold cross-validation on the training set with the area under the receiver operating characteristic curve (AUC) being the main evaluation measure.

The random forest (RF) model was chosen for further development in light of its cross-validated discriminative performance, and the hyperparameters of this model were optimized through a combination of stepwise tuning and Bayesian optimization so as to maximize the cross-validated AUC.

### Feature Selection and Model Parsimony

In order to cut down on model intricacy and boost interpretability, a method based on SHAP was utilized for feature selection. The significance of features was quantified through SHAP, which gauges the contribution of every feature to model forecasts by computing SHAP figures within the training data in a 10-fold cross validation.

In every cross-validation fold, SHAP values were calculated for the relevant validation subset and the significance of features was summarized via mean absolute SHAP values. After that the rankings of features were combined across all folds to get a stable estimation of overall feature importance.

In order to find a concise set of features, features were added one after another to the model in accordance with the aggregated SHAP importance ranking beginning with the most crucial feature and at every step, the model’s performance was assessed by means of 10-fold cross-validation and the connection between the quantity of incorporated features and the cross-validated AUC was investigated.

The simplified model was specified as the one attaining the top cross-validated AUC having the least quantity of features and this approach equalized predictive capability and model straightforwardness whilst maintaining the most useful variables.

### Model Evaluation and Threshold Determination

After selecting the features, the simplified random forest (RF) model was evaluated within a 10-fold cross-validation framework based on the training data and its discriminative ability was mainly measured by the AUC.

In the 10-fold cross-validation process, an ideal probability threshold was worked out according to the Youden index and with this threshold obtained from cross validation[16]. The final simplified model along with its matching probability threshold was subsequently utilized for performance assessment on the independent testing set and apart from AUC, threshold-reliant classification metrics such as accuracy, precision, sensitivity, specificity, F1 score, and geometric mean (G-mean) were computed.

To evaluate the sturdiness, the whole procedure of data division, model training, and testing was repeated with 10 distinct random seeds and the model performance figures were compared among these re-run experiments.

### Model Interpretability and Nonlinear Effect Analysis

The interpretability of the model was evaluated by means of SHAP so as to measure how much each individual feature contributed to the predictions made by the simplified RF model. In order to get stable estimates of the overall importance of different features, SHAP values were computed on the independent testing sets within 10 repeated random data splits and the importance of features was summed up by taking the average of the absolute SHAP values collected from these testing sets.

To probe into the nonlinear connections between key continuous features and model predictions, SHAP dependence plots were created based on the predictions from the testing sets, which depicted the association between feature values and their respective SHAP values. And locally weighted smoothing was utilized to picture overall trends and identify potential inflection points characterized by changes in slope, which were determined at the points corresponding to the maximum slope of the smoothed curves.

To further verify the nonlinear relationships that had been observed, partial dependence plots (PDPs) were constructed for the same features by employing the trained simplified model. PDPs estimated how much a particular feature influenced the predicted results when considering the average influence of other characteristics across their distributions, then the similarity between SHAP dependence plots and PDPs was utilized to back up the stability of the detected nonlinear patterns.

## Results

### Study Population and Feature Overview

A total of 770 eyes of 594 patients with high myopia were included into the analysis, among which 458 eyes didn’t have cataracts while 312 eyes have cataracts. Demographic traits such as age and sex, ocular biometric factors, systemic laboratory factors, and calculated composite indexes were considered as potential features. After discarding features with over 30% missing data, 56 features were kept for model development. The distribution of missing values within the retained features (derived composite indexes excluded) can be seen in ***Supplementary Figure S1***. The data distribution was shown in **Table 1**.

**Table 1.**
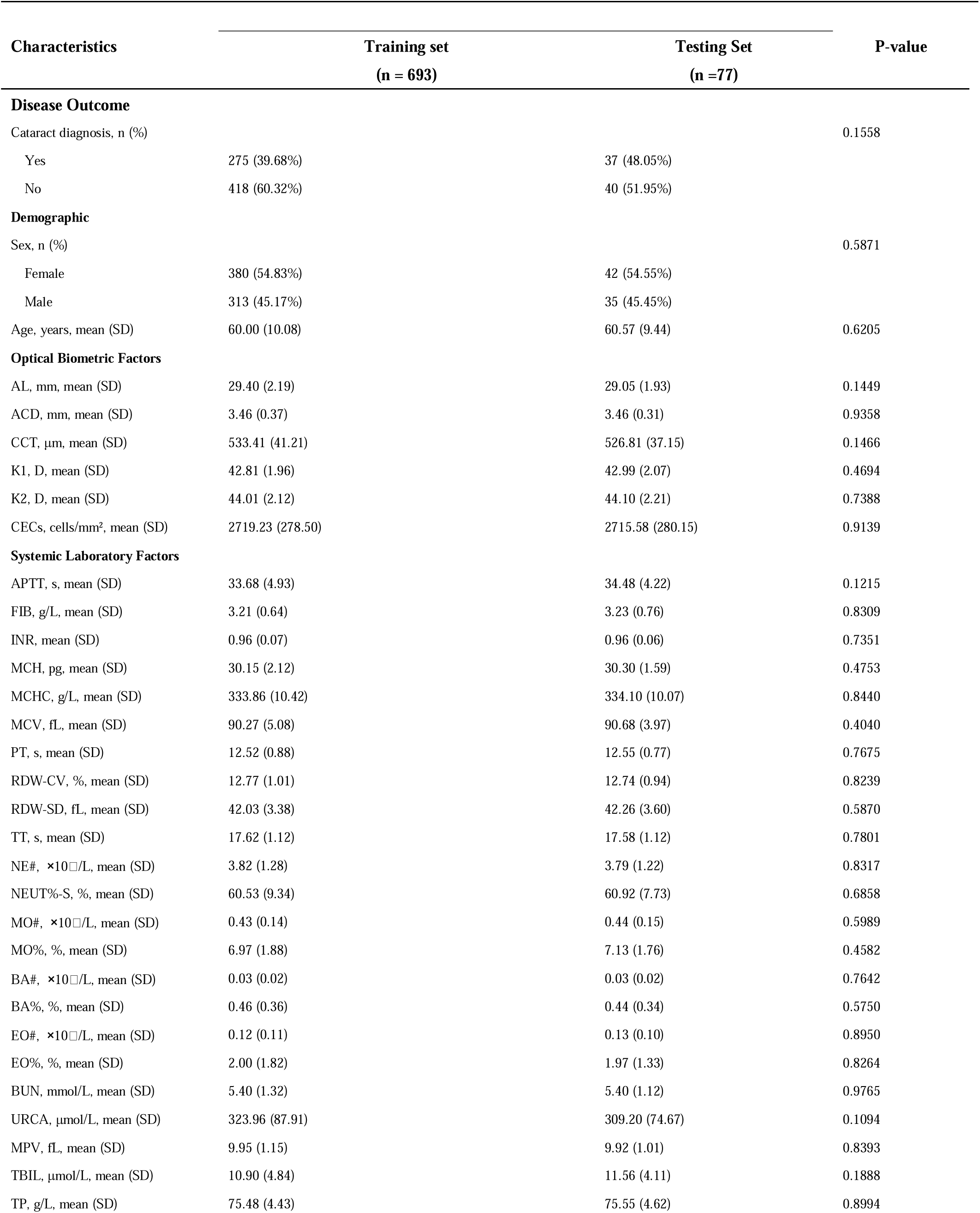
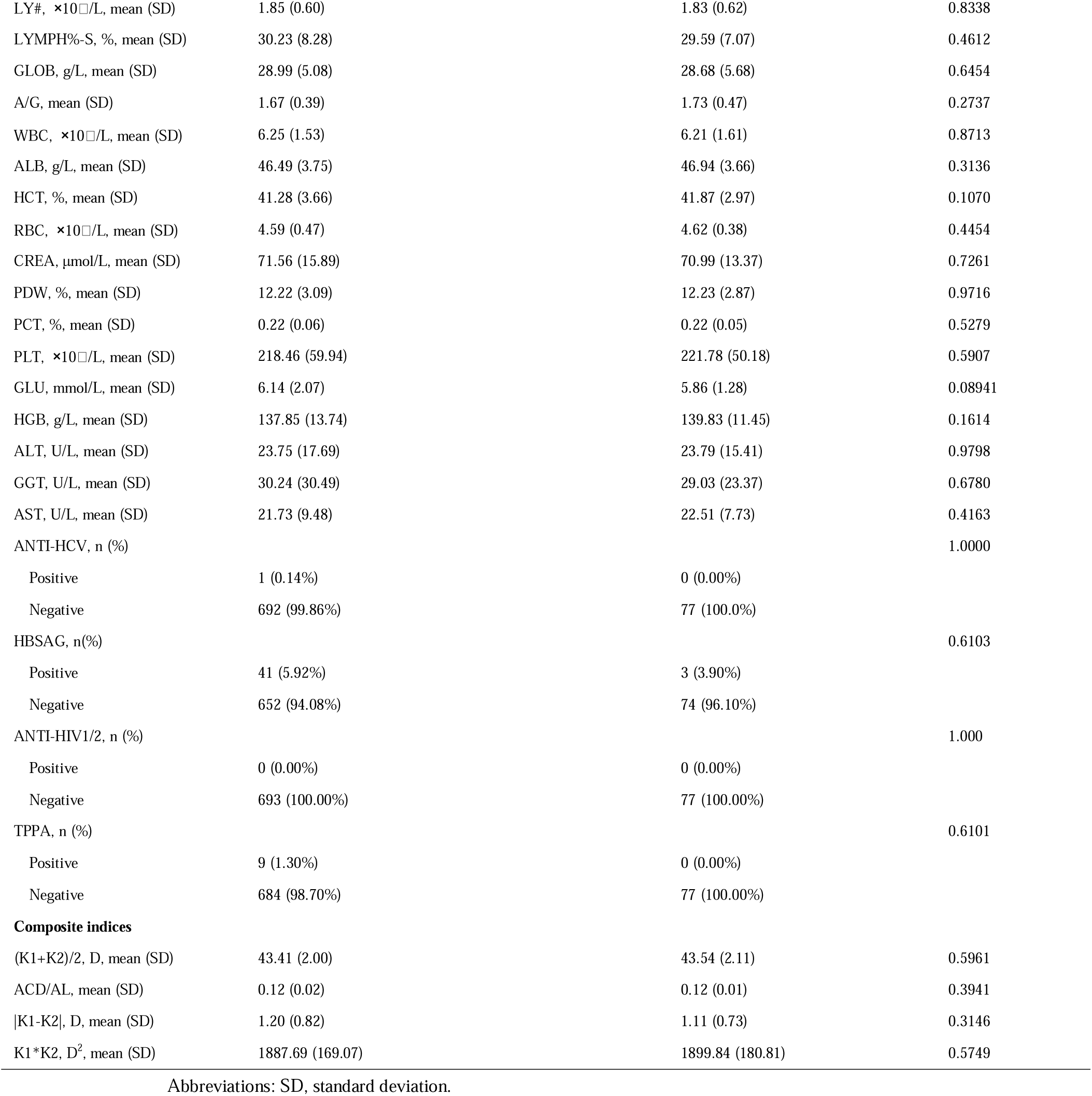
Baseline characteristics of the training and testing set.

### Model Comparison and Selection

Numerous machine learning models were evaluated via 10-fold cross-validation utilizing the training data. Comparative performance of these models, which was assessed by the AUC, is encapsulated in ***Supplementary Figure S2A and S2B***. Among all the evaluated models, the RF model exhibited the top mean cross-validated AUC, so it was consequently chosen for further model training.

The RF was later optimized through a two-stage tuning approach. In the initial phase, stepwise tuning was carried out to determine a rational range of hyperparameters and then Bayesian optimization was utilized to further fine-tune the model parameters (as shown in ***Supplementary Figure S3***). After the hyperparameters were optimized, the mean AUC derived from 10-fold cross-validation went up by 1.45% in comparison to that of the untuned model, attaining an average AUC of 0.760 (95%CI: [0.734,0.786]), as shown in ***Supplementary Figure S2C***. And the optimized hyperparameter settings of the RF model are listed in ***Supplementary Table S2***. The optimized RF model was employed for all the subsequent analyses.

### Feature Selection and Model Parsimony

A SHAP-based method was utilized for feature selection within a 10-fold cross-validation setup for each fold. SHAP values were computed and then the feature importance rankings were combined across different folds, the outcomes of the cross-validated SHAP examination can be found in ***Supplementary Figure S4***.

To find an ideal and concise set of features, features were added one after another to the RF model based on their SHAP importance rank and the model’s performance was assessed at every step through 10-fold cross-validation, as shown in **Figure 2A**, the cross-validated AUC went up when top-ranked features were included and attained its peak when 17 features were incorporated, resulting in a mean AUC of 0.773 (95%CI: [0.748, 0.797]).

**Figure 2.**
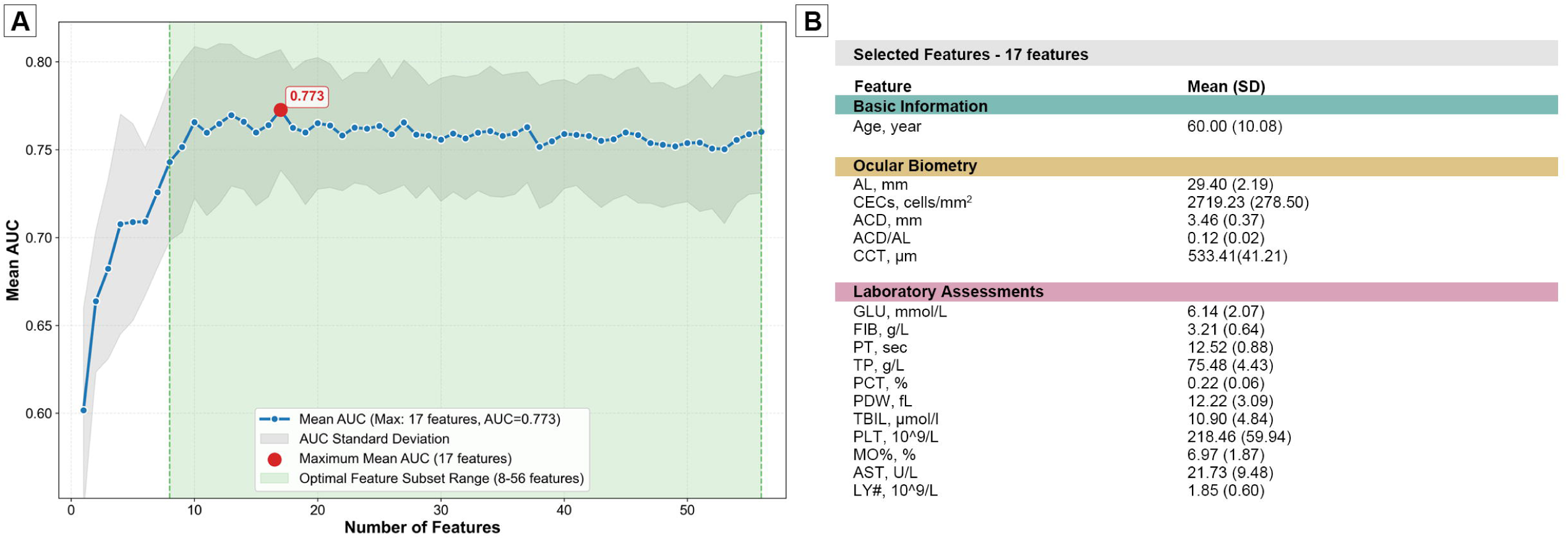
Feature selection and identification of a simplified RF model. (A) The connection between the quantity of incorporated features and cross-validated model performance: Characteristics were progressively added in line with their SHAP significance hierarchy obtained from 10-fold cross-validation, and the average area under the receiver operating characteristic curve (AUC) attained its peak when 17 characteristics were integrated, achieving a cross - validated AUC of 0.773 (95%CI: [0.748, 0.797]); (B) Makeup of the chosen 17-feature group forming the simplified model.

The composition of the selected 17-feature subset is shown in **Figure 2B**. This simplified feature set was therefore adopted for subsequent model optimization, evaluation, and interpretability analyses.

### Feature Importance on Independent Testing Sets

In order to depict the feature importance within the final simplified model, SHAP analysis was carried out on the independent testing sets over ten times of repeated random data divisions. And the feature importance was summarized by means of average absolute SHAP values which were accumulated across these testing sets.

The global SHAP importance ranking of the top 15 characteristics is shown in **Figure 3A and Figure 3B**. Age and AL were constantly considered among the most influential features throughout the testing sets and then came ACD, CECs and ACD/AL. The relative significance of the leading features demonstrated high consistency across random data divisions (**Figure 3C and Figure 3D**), which signified that the feature contributions to model performances were stable.

**Figure 3.**
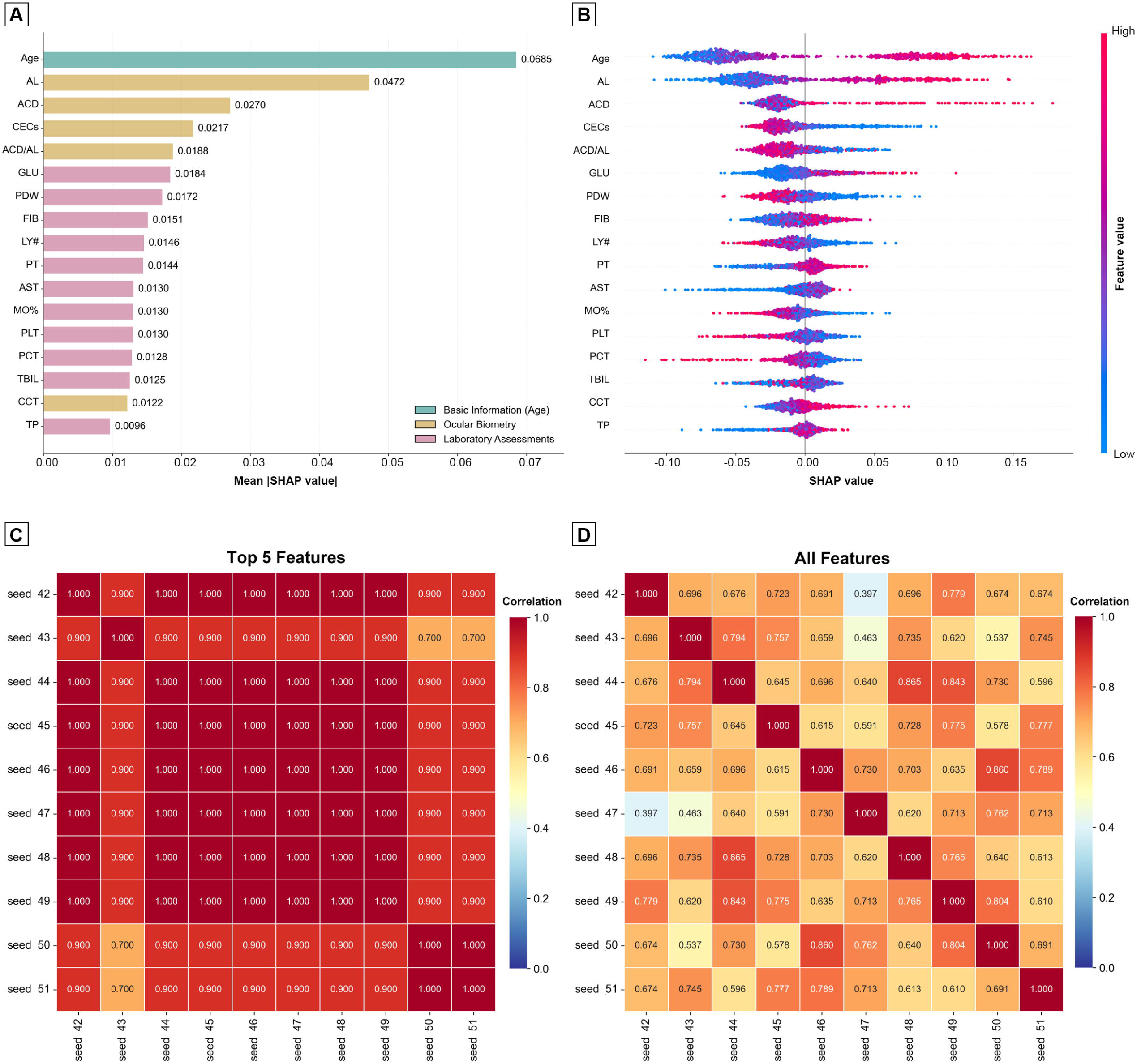
The SHAP feature importance was obtained from independent testing sets and the SHAP values were computed on these independent testing sets over 10 repeated random data partitions before being aggregated to evaluate the overall feature significance. (A) presents a bar graph which displays the top 15 features sorted according to their mean absolute SHAP values across the testing sets; (B) shows a SHAP beeswarm diagram that depicts the distribution of SHAP values for these same top 15 features indicating both the size and direction of their influences on model predictions; (C) demonstrates the consistency regarding the top five feature importance rankings across the 10 testing sets which originated from diverse random seeds and part; (D) illustrates the consistency of SHAP feature importance rankings for all features throughout the 10 testing sets.

These results highlight a small set of dominant features driving model predictions and provide the basis for subsequent analyses of nonlinear effects and clinically relevant patterns.

### Nonlinear Effects and Threshold Identification of Key Features

To further delve into how crucial continuous characteristics impacted model forecasts, SHAP dependence analyses were carried out on the independent test sets throughout 10 repeated random data divisions and for every characteristic, the SHAP values from the test sets were combined to depict nonlinear patterns and spot constant trends.

#### Nonlinear Effects and Thresholds of Age and Axial Length

SHAP dependence diagrams showed that SHAP values monotonically rose as age increased, which signified that age made an ever-growing contribution to the forecasted cataract risk (**Figure 4A and Figure 4B**). And the connection was non-linear with a distinct alteration in the slope. In each of the ten testing groups, for every division, the point where the steepest slope occurred was found and on average, the inflection point was noticed at 65.75 (95%CI: [63.72, 67.79]) years.

**Figure 4.**
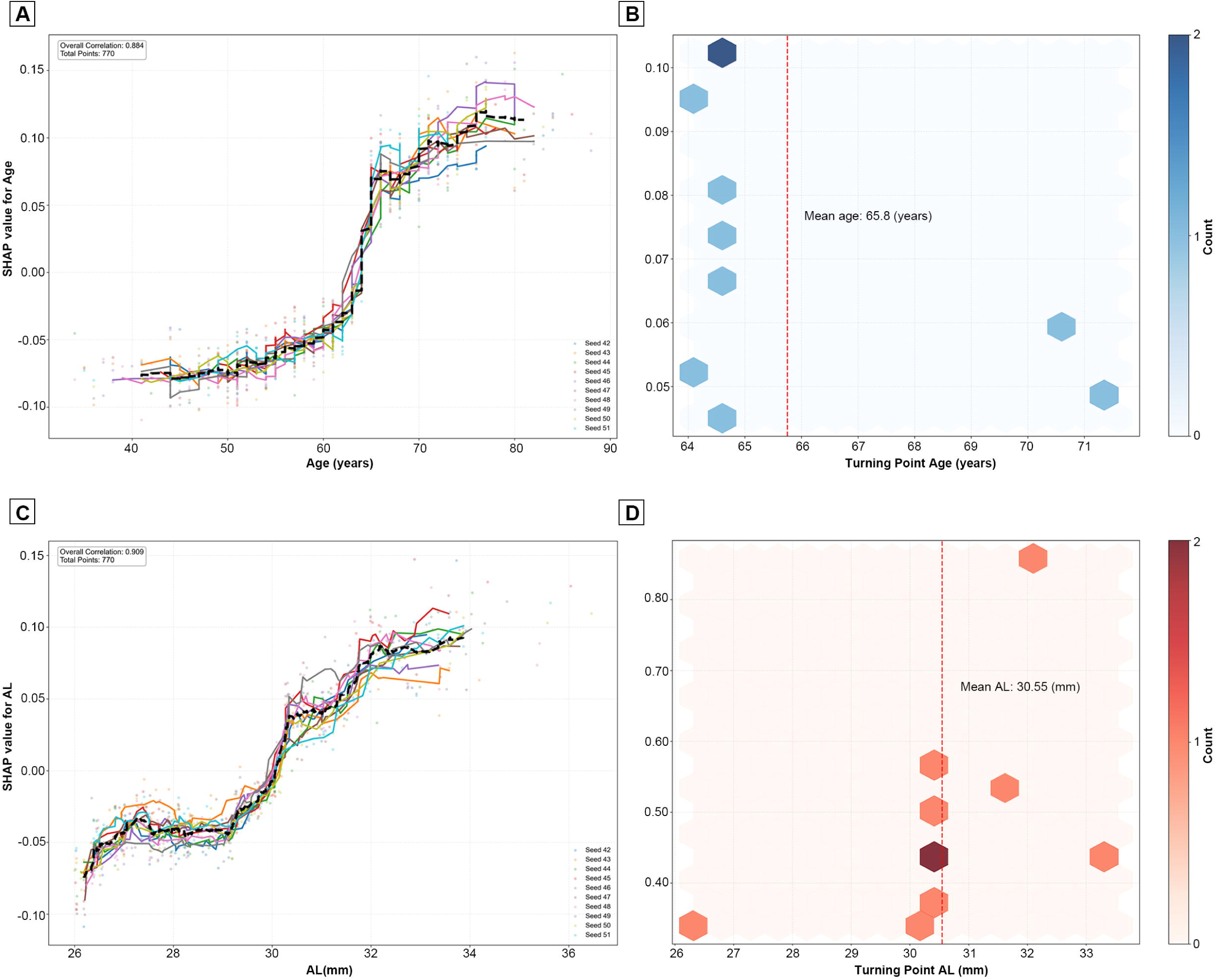
Nonlinear threshold effects of age and axial length disclosed by SHAP dependence examinations. (A) SHAP dependence diagrams for age throughout the independent testing groups where every curve stood for the locally smoothed connection between age and SHAP figures obtained from one testing set demonstrating uniform nonlinear tendencies across the ten data divisions; (B) A schematic overview of the age-associated turning points detected in each testing group matching the points of the steepest gradient in the SHAP dependence curves with the average turning point among testing groups marked; (C) SHAP dependence graphs for axial length (AL) over the independent testing groups in which each curve denoted one testing group indicating a nonlinear rise in SHAP values as AL increased; (D) A schematic synopsis of the AL-related turning points found in each testing group with the average turning point across testing groups shown.

Similarly, AL had a non-linear connection with SHAP values where greater AL was linked to higher SHAP values and a increased contribution in model performances (**Figure 4C and Figure 4D**). An obvious acceleration of SHAP values could be seen at longer axial lengths and the average inflection point related to the steepest slope across the testing sets was 30.55 (95% CI: [29.22, 31.88]) mm showing a threshold beyond which the contribution of AL grew much faster.

#### U-Shaped Association of ACD/AL With Model Predictions

In dependency analyses, ACD/AL had a U-shaped association with SHAP values as seen in **Figure 5A**, with SHAP values being greater at both the lower and upper extremes of ACD/AL and intermediate values linked to lower SHAP values, which indicated a non - monotonic contribution pattern.

**Figure 5.**
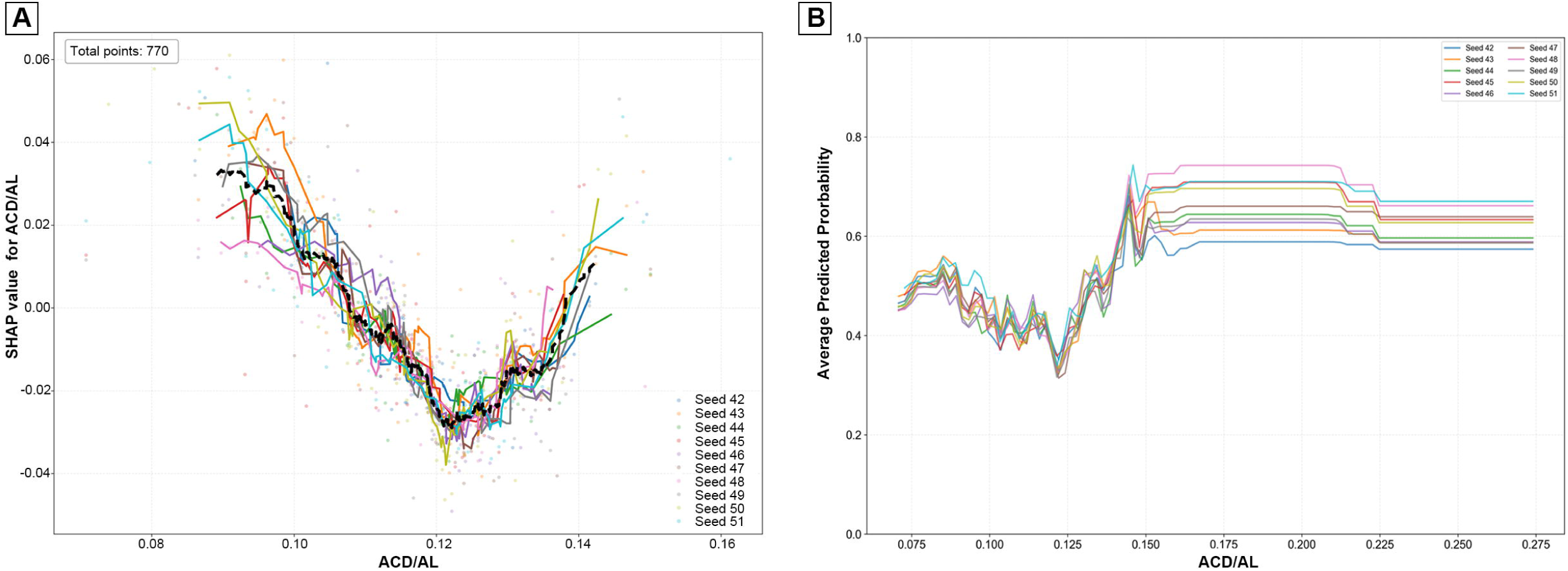
U-shaped connection of the ratio of anterior chamber depth to axial length (ACD/AL) with model forecasts. (A) The SHAP dependence diagram showed a U - shaped link between the ACD/AL ratio and the SHAP figures obtained from the separate testing groups, with greater SHAP figures noticed at both the lower and upper ends of ACD/AL; (B) The partial dependence plot (PDP) for ACD/AL indicated a similar U-shaped pattern and supported the non-linear relation seen in the SHAP dependence examination.

PDPs built for ACD/AL displayed a steady U-shaped tendency (**Figure 5B**), which was in line with the pattern found in the SHAP dependence analysis and the agreement between SHAP dependence plots and PDPs across testing sets indicated that this non-linear connection was robust.

### Model Performance and Threshold Determination

The performance of the final simplified RF model was further assessed on independent testing sets over ten repeated random data divisions and its discriminant performance stayed steady on the testing sets, with a mean AUC of 0.762 (95%CI: [0.731, 0.794]) which supported the model’s robustness and detailed performance figures for every testing set were given in ***Supplementary Table S3***.

To figure out an operating threshold for binary classification, the ideal probability cut-off was chosen according to the Youden index which came from 10-fold cross-validation on the training data and the highest Youden index was 0.411, matching an ideal probability threshold of 0.423 and the connection between the Youden index and classification threshold could be seen in ***Supplementary Figure S5***.

## Discussion

In this research, we utilized an interpretable machine learning method to probe into age, sex, biometric factors, and systemic laboratory factors linked to cataract in highly myopic eyes. By integrating robust model development with feature importance and non-linear pattern analyses, we found a simplified model which showed relatively steady discriminative ability while providing findings that could be interpreted clinically and crucially. The outcomes highlighted a limited number of predominant features and distinctive non-linear patterns associated with cataract in highly myopic eyes.

Across every feature, age and AL continuously demonstrated the most robust connections to cataract within independent testing sets and this finding was in line with long-established clinical knowledge that as one grows older, the likelihood of developing cataract rises and in highly myopic eyes, while excessive elongation of the axis also contributes to it, though previous studies were generally based on small sample sizes or did not examine the nonlinear relationship between age, AL, and high myopic cataract[17–19].

Our study discovered that, besides their general significance, both age and AL exhibited nonlinear relationships with the presence of cataracts as their influences grew step by step with greater values and had clear inflection points after which the strength of the relationship became more evident, the average inflection points were around 65.8 years for age and 30.55 (95% CI: [29.22, 31.88]) mm for axial length and such values were constantly seen in repeated testing sets. Clinically, the growing role of age probably mirrored cumulative lens-aging processes like long-term exposure to oxidative stress, gradual changes in protein structure, and reduced cellular repair ability[20, 21]. And likewise excessive axial elongation might be linked to abnormal tension in the zonular apparatus in very myopic eyes which could affect lens position and disrupt normal lens metabolism[22], and these structural and metabolic changes might lead to greater vulnerability to cataract development in eyes with extreme AL. Although these explanations were still speculative, they were in line with existing clinical and experimental findings.

Besides age and AL, ACD/AL had a U-shaped connection with cataract, with eyes having either a rather low or high ACD/AL ratio showing more pronounced connections while intermediate values were linked to less significant contributions and this pattern was constantly seen across different analytical methods. Clinically speaking, a low ACD/AL ratio might mean that the anterior chamber is relatively shallow in an elongated eye, which could possibly influence lens positioning, iris-lens interaction, or aqueous humor dynamics[23–25]. While a high ACD/AL ratio could show that the anterior segment has become abnormally long compared to the posterior segment, which might be related to changes in zonular tension or lens stability[26], indicating that combined measures like ACD/AL may more accurately represent the overall ocular shape and its relationship with cataract in highly myopic eyes when compared to single biometric measurements alone.

One significant advantage of this research was the constancy of results throughout various data divisions and analysis procedures, as feature choice and the evaluation of their significance were carried out meticulously with the training and testing data clearly distinguished, and the steadiness of crucial connections during multiple testing sets strengthened the reliability of the noticed patterns, while it was crucial that the application of understandable analysis instruments made it possible to see how single features associated with the existence of cataracts over their entire scope, which simplified clinical understanding instead of just focusing on predictive modeling.

Clinically speaking, the identification of non-linear and threshold-like associations might have ramifications for evaluating and monitoring patients. Instead of presuming a uniform, linear connection between ocular biometric factors and cataracts, these results imply that the strength of this association could vary beyond specific ranges of age or AL so such non-linear tendencies might mirror fundamental biological shifts[27], such as age-related modifications in lens metabolism and AL-associated alterations in eye biomechanics and zonular tension, which altogether could impact lens equilibrium. Crucially, these figures shouldn’t be regarded as diagnostic or management cut-offs. Instead, they are data-based patterns that could assist in sharpening clinical understanding of stage-dependent structural and metabolic changes in highly myopic eyes and direct future long-term research aiming to clarify the mechanisms connecting ocular anatomy and cataract development.

## Conclusion

In brief, this research showed that an interpretable, data-based method was able to find strong and clinically significant associations between ocular features and highly myopic cataracts, underlined the important parts played by age, AL, and composite biometric indices like ACD/AL, which emphasized the close relationship between ocular biometrics and highly myopic cataracts. Notably, we further observed nonlinear associations of age and AL with highly myopic cataract, suggesting that potential inflection points may exist. Together, these findings indicate that ocular biometric factors may reflect the fundamental structural and metabolic changes in highly myopic eyes related to cataract development, thus providing a pattern for future investigations focused on clarifying disease processes, making the description of phenotypes more precise, and enhancing the clinical knowledge of highly myopic cataracts.

## Data Availability

All data produced in the present study are available upon reasonable request to the authors

## Authors contributions

K.S., W.H. participated in searching databases and preparing data. K.S. and W.H. analyzed the data and prepared the figures and tables. K.S. and Q.D. prepared the original draft of the manuscript. Q.D. provided methodological guidance. X.Z., L.G., B.W. reviewed and edited the manuscript. X.Z., L.G., B.W. and R.E. conceived and supervised the project. All authors have read and agreed to the published version of manuscript.

## Consent for publication

Not applicable

## Supplementary information

This article contains additional online-only material. The following should appear online-only: Supplementary Fig S1-5, and Table S1-3.

