## Supplementary Figure 1-5 for "Interpretable machine-learning model for cataract associated factors identifying in patients with high myopia"

**Supplementary Figure S1. Missing data distribution of included features**

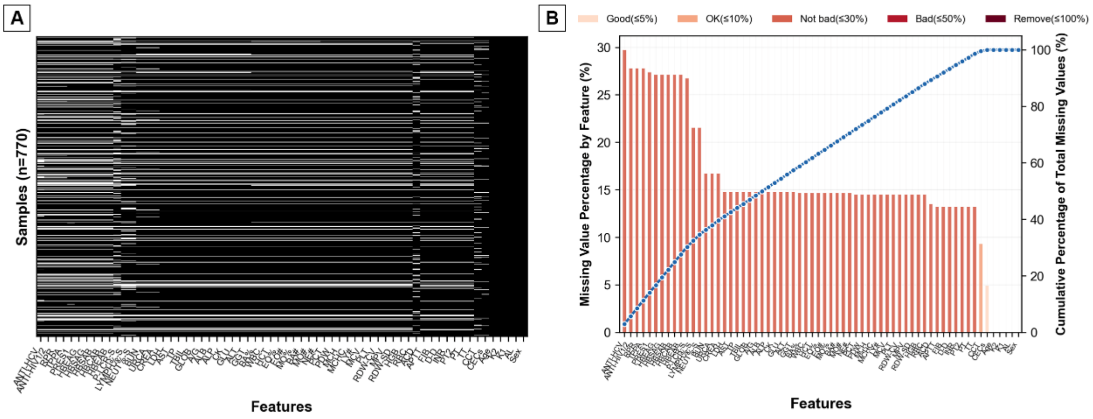


(A) Heatmap illustrating the pattern of missing values across the 52 retained original features (excluding derived composite indices) prior to imputation. Each column represents a feature, and each row represents an eye.

(B) Pareto plot showing the proportion of missing values for each feature, ranked in descending order.

**Supplementary Figure S2. Comparison of machine learning models and hyperparameter optimization of the random forest model.**

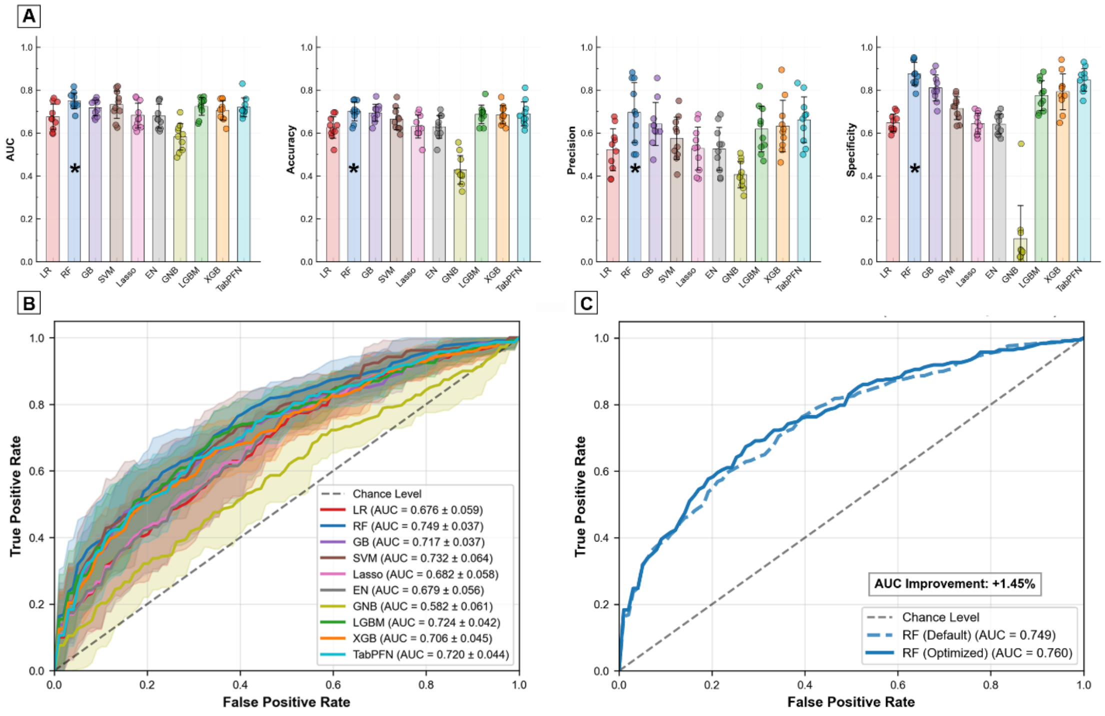


(A) Bar plots showing the mean performance of the 10 evaluated machine learning models based on 10-fold cross-validation on the training data, including area under the receiver operating characteristic curve (AUC), accuracy, precision, and specificity; (B) Receiver operating characteristic (ROC) curves of the 10 machine learning models obtained from 10-fold cross-validation on the training data; (C) ROC curves of the random forest model before and after hyperparameter optimization, illustrating the improvement in discriminative performance following the two-stage tuning strategy.

**Supplementary Figure S3. Two-stage hyperparameter optimization of the random forest model.**

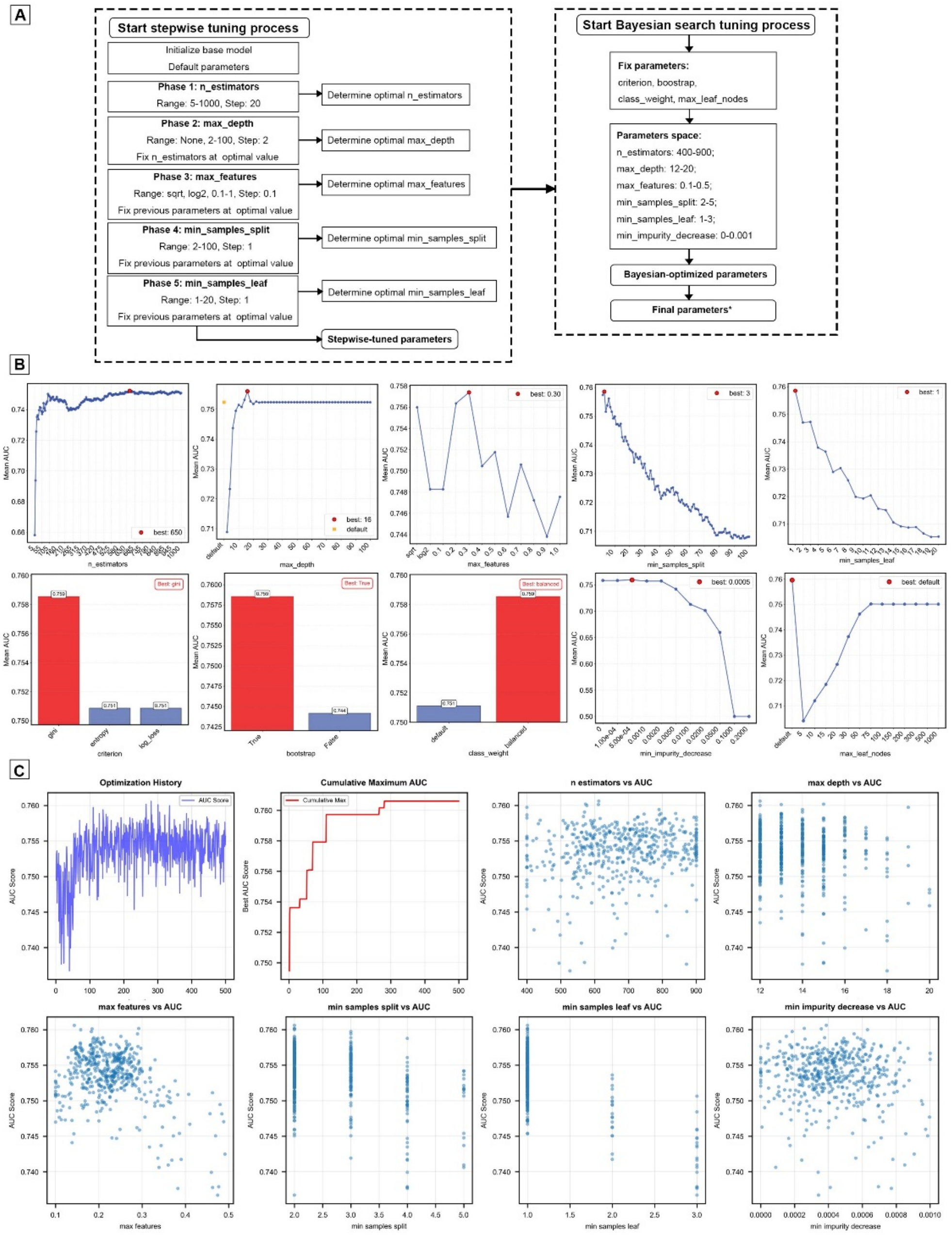


(A) Schematic illustration of the two-stage tuning strategy, including stepwise tuning to identify a reasonable hyperparameter range followed by Bayesian optimization to further refine model parameters; (B) Results of stepwise tuning, showing the performance of different hyperparameter configurations evaluated during the initial tuning stage; (C) Results of Bayesian optimization, illustrating the explored hyperparameter space and the corresponding model performance during the optimization process.

**Supplementary Figure S4. Cross-validated SHAP analysis of feature importance.**

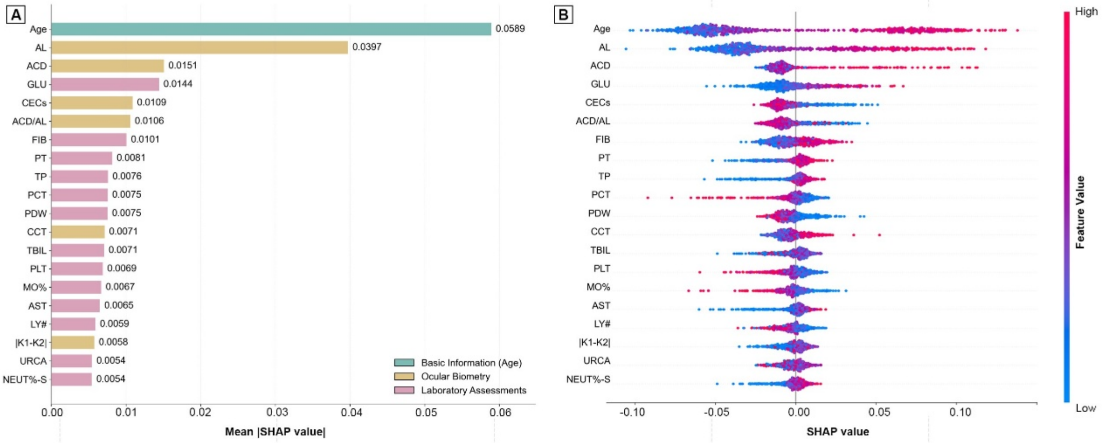


SHAP values were calculated within a 10-fold cross-validation framework on the training data and aggregated across folds to rank feature importance.

(A) Bar plot showing the top 20 features ranked by mean absolute SHAP values, summarizing their overall contribution to model predictions; (B) SHAP beeswarm plot illustrating the distribution of SHAP values for the same top 20 features across cross-validation folds, reflecting both the magnitude and direction of feature contributions.

**Supplementary Figure S5. Threshold determination and decision curve analysis of the**

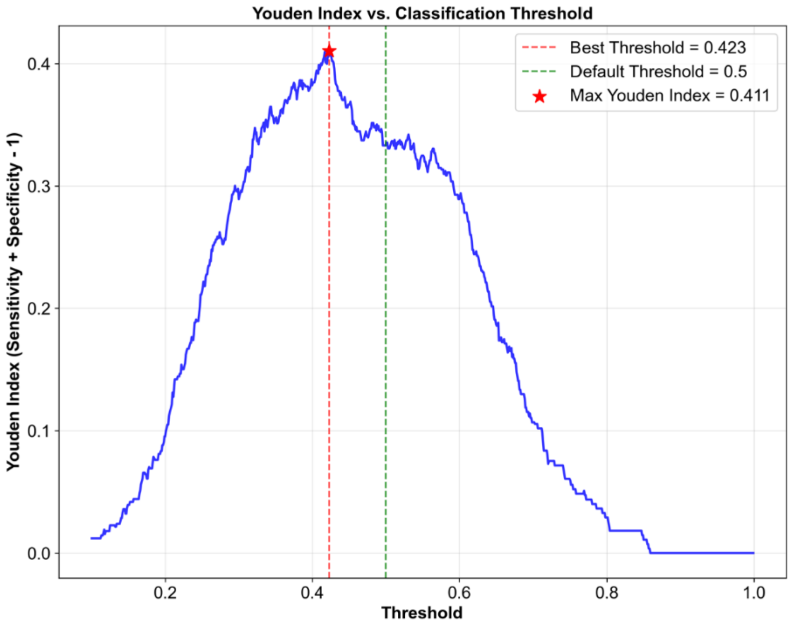


**parsimonious random forest model.**

Relationship between the Youden index and classification probability threshold derived from 10-fold cross-validation on the training data. The optimal threshold corresponding to the maximum Youden index is indicated.
