## Supplementary Table 1-3 for "Interpretable machine-learning model for cataract associated factors identifying in patients with high myopia"

**Supplementary Table S1. List and definitions of features included in model development.**

| Category | Feature name | Description | Unit |
|---|---|---|---|
| Demographic | Age | Age at presentation | years |
| Demographic | Sex | Biological sex | - |
| Ocular biometry | AL | Axial length | mm |
| Ocular biometry | K1 | Flat keratometry | D |
| Ocular biometry | K2 | Steep keratometry | D |
| Ocular biometry | ACD | Anterior chamber depth | mm |
| Ocular biometry | CCT | Central corneal thickness | μm |
| Ocular biometry | CECs | Corneal endothelial cell density | cells/mm² |
| Laboratory | APTT | Activated partial thromboplastin time | s |
| Laboratory | PT | Prothrombin time | s |
| Laboratory | INR | International normalized ratio | - |
| Laboratory | TT | Thrombin time | s |
| Laboratory | FIB | Fibrinogen | g/L |
| Laboratory | WBC | White blood cell count | $\times 10^9$/L |
| Laboratory | NE# | Neutrophil count | $\times 10^9$/L |
| Laboratory | NEUT%-S | Neutrophil percentage | % |
| Laboratory | LY# | Lymphocyte count | $\times 10^9$/L |
| Laboratory | LYMPH%-S | Lymphocyte percentage | % |
| Laboratory | MO# | Monocyte count | $\times 10^9$/L |
| Laboratory | MO% | Monocyte percentage | % |
| Laboratory | EO# | Eosinophil count | $\times 10^9$/L |
| Laboratory | EO% | Eosinophil percentage | % |
| Laboratory | BA# | Basophil count | $\times 10^9$/L |
| Laboratory | BA% | Basophil percentage | % |
| Laboratory | RBC | Red blood cell count | $\times 10^{12}$/L |
| Laboratory | HGB | Hemoglobin | g/L |
| Laboratory | HCT | Hematocrit | % |
| Laboratory | MCV | Mean corpuscular volume | fL |
| Laboratory | MCH | Mean corpuscular hemoglobin | pg |
| Laboratory | MCHC | Mean corpuscular hemoglobin concentration | g/L |
| Laboratory | RDW-CV | Red cell distribution | % |

| | | | |
|---|---|---|---|
| | | width (coefficient of variation) | |
| Laboratory | RDW-SD | Red cell distribution width (standard deviation) | fL |
| Laboratory | PLT | Platelet count | $\times 10^9$/L |
| Laboratory | MPV | Mean platelet volume | fL |
| Laboratory | PDW | Platelet distribution width | % |
| Laboratory | PCT | Platelet | % |
| Laboratory | GLU | Blood glucose | mmol/L |
| Laboratory | BUN | Blood urea nitrogen | mmol/L |
| Laboratory | CREA | Creatinine | μmol/L |
| Laboratory | URCA | Uric acid | μmol/L |
| Laboratory | TP | Total protein | g/L |
| Laboratory | ALB | Albumin | g/L |
| Laboratory | GLOB | Globulin | g/L |
| Laboratory | A/G | Albumin-to-globulin ratio | - |
| Laboratory | TBIL | Total bilirubin | μmol/L |
| Laboratory | ALT | Alanine aminotransferase | U/L |
| Laboratory | AST | Aspartate aminotransferase | U/L |
| Laboratory | GGT | Gamma-glutamyl transferase | U/L |
| Laboratory | HBSAG | Hepatitis B surface antigen | - |
| Laboratory | ANTI-HCV | Hepatitis C virus antibody | - |
| Laboratory | ANTI-HIV1/2 | Human immunodeficiency virus antibody | - |
| Laboratory | TPPA | Treponema pallidum particle agglutination assay | - |
| Composite index | (K1+K2)/2 | Mean keratometry | D |
| Composite index | ACD/AL | Ratio of anterior chamber depth to axial length | - |
| Composite index | \|K1−K2\| | Absolute keratometric astigmatism | D |
| Composite index | K1×K2 | Product of keratometry | $D^2$ |

| | | |
|---|---|---|
| | | values |

Ocular biometric factors were measured using an optical biometer (IOLMaster 500 and 700, Carl Zeiss, Germany). Units and reference ranges followed institutional laboratory standards.

**Supplementary Table S2. Hyperparameter tuning of the random forest model**

| Hyperparameter | Default Parameters | Stepwise-Tuned Parameters | Bayesian-Optimized Parameters |
|---|---|---|---|
| **Number of trees** | | | |
| n_estimators | 100 | 650 | 612 |
| **Tree complexity control** | | | |
| max_depth | None | 16 | 12 |
| max_leaf_nodes | None | None | None |
| min_impurity_decrease | 0.0 | 0.0005 | 0.0003872517226561781 |
| **Node splitting criteria** | | | |
| criterion | 'gini' | 'gini' | |
| **Features for split** | | | |
| max_features | 'sqrt' | 0.3 | 0.14333272138401731 |
| Sample requirements | | | |
| min_samples_split | 2 | 3 | 2 |
| min_samples_leaf | 1 | 1 | 1 |
| **Bootstrapping** | | | |
| bootstrap | True | True | True |
| Class balance | | | |
| class_weight | 'balanced' | 'balanced' | 'balanced' |

**Supplementary Table S3. Performance metrics of the parsimonious random forest model across independent testing sets.**

| Seeds | AUC | Accuracy | Precision | Sensitivity | F1_score | Specificity | G_mean |
|---|---|---|---|---|---|---|---|
| 42 | 0.767 | 0.701 | 0.750 | 0.568 | 0.646 | 0.825 | 0.684 |
| 43 | 0.764 | 0.688 | 0.610 | 0.758 | 0.676 | 0.636 | 0.694 |
| 44 | 0.742 | 0.688 | 0.656 | 0.618 | 0.636 | 0.744 | 0.678 |
| 45 | 0.791 | 0.727 | 0.707 | 0.763 | 0.734 | 0.692 | 0.727 |
| 46 | 0.786 | 0.727 | 0.613 | 0.679 | 0.644 | 0.755 | 0.716 |
| 47 | 0.808 | 0.740 | 0.771 | 0.692 | 0.730 | 0.789 | 0.739 |
| 48 | 0.656 | 0.584 | 0.500 | 0.469 | 0.484 | 0.667 | 0.559 |
| 49 | 0.750 | 0.623 | 0.632 | 0.615 | 0.623 | 0.632 | 0.623 |
| 50 | 0.805 | 0.714 | 0.632 | 0.750 | 0.686 | 0.689 | 0.719 |
| 51 | 0.746 | 0.662 | 0.419 | 0.619 | 0.500 | 0.679 | 0.648 |
